# The impact of the COVID-19 pandemic on influenza, respiratory syncytial virus, and other seasonal respiratory virus circulation in Canada

**DOI:** 10.1101/2021.04.15.21255591

**Authors:** HE Groves, P Piché-Renaud, A Peci, DS Farrar, S Buckrell, C Bancej, C Sevenhuysen, A Campigotto, JB Gubbay, SK Morris

## Abstract

**Background:** The ongoing coronavirus disease 2019 (COVID-19) pandemic has resulted in implementation of public health measures worldwide to mitigate disease spread, including; travel restrictions, lockdowns, messaging on handwashing, use of face coverings and physical distancing. As the pandemic progresses, exceptional decreases in seasonal respiratory viruses are increasingly reported. We aimed to evaluate the impact of the pandemic on circulation of influenza, respiratory syncytial virus and other seasonal respiratory viruses in Canada.

**Methods:** Epidemiologic data were obtained from the Canadian Respiratory Virus Detection Surveillance System. Weekly data from the week ending 30^th^ August 2014 until the week ending the 13^th^ February 2021 were analysed. We compared trends in laboratory detection and test volumes during the 2020/2021 influenza season with baseline pre-pandemic seasons from 2014 to 2019.

**Findings:** We observed a dramatically lower percentage of tests positive for all seasonal respiratory viruses during 2020-2021 compared to baseline. For influenza A and B the percent positive decreased to 0·0017 and 0·0061 times that of baseline respectively and for RSV, the percent positive dropped to 0·0145 times that of baseline. Ongoing detection of enterovirus/rhinovirus occurred, with regional variation in the epidemic patterns and intensity.

**Interpretation:** We report an effective absence of the annual seasonal epidemic of most seasonal respiratory viruses in 2020/2021. This dramatic decrease is likely related to implementation of multi-layered public health measures during the pandemic. The impact of such measures may have relevance for public health practice in mitigating seasonal respiratory virus epidemics and for informing responses to future respiratory virus pandemics.

**Funding:** No additional funding source was required for this study.

**Research in context:** *Evidence before this study:* We searched PubMed, preprint servers and country-specific public health rapid communications to identify surveillance and epidemiological studies on influenza, respiratory syncytial virus and other seasonal respiratory virus detection during the COVID-19 pandemic. A number of regional and national studies were identified worldwide. The majority of these studies focus on influenza epidemiology and all studies show consistent decreases in circulation of seasonal non-SARS-CoV-2 respiratory viruses. One previous study on the impact of non-pharmaceutical interventions on laboratory detections of influenza A and B in Canada included data for the 2019/2020 influenza season. Another recent study examined the effect of seasonal respiratory virus transmission on COVID-19 syndromic surveillance in the province of Ontario, Canada. No previous Canada-wide study has described the epidemiology of influenza, respiratory syncytial virus and other seasonal respiratory virus detection during the 2020/2021 influenza season.

*Added value of this study:* The Canadian Respiratory Virus Detection Surveillance System provides weekly respiratory virus detection reports from sentinel laboratories across Canada for influenza, respiratory syncytial virus, parainfluenza viruses, adenovirus, human metapneumovirus, enterovirus/rhinovirus and seasonal coronaviruses. Data have been collected continuously since 2004. Analysis of this dataset provides a comprehensive assessment of the impact of the COVID-19 pandemic on circulation of seasonal respiratory viruses in Canada and analysis of data from the Canadian Public Health Infobase on COVID-19 allowed comparison of SARS-CoV-2 epidemiology. This is the first country-wide study in the Northern hemisphere to describe the concurrent epidemiology of all major seasonal respiratory viruses and SARS-CoV-2 during the 2020/2021 influenza season.

*Implications of all the available evidence:* The effective absence of the annual seasonal epidemic for most non-SARS-CoV-2 respiratory viruses in 2020/2021 has important public health implications for informing ongoing and future responses to respiratory virus epidemics and pandemics.

## Introduction

The emergence of coronavirus disease 2019 (COVID-19) has resulted in an unprecedented global pandemic leading to significant morbidity and mortality, particularly among older and vulnerable adult populations.^1^ It has been more than one year since the pandemic first began and in this time, policy makers worldwide have implemented stringent mitigation efforts to reduce transmission of severe acute respiratory syndrome virus 2 (SARS-CoV-2). These measures have included the implementation of local and international travel restrictions; use of targeted lockdowns including stay-at-home orders and school closures; universal guidance on handwashing, physical distancing and, use of face coverings.^2,3^ In Canada, restrictions on international travel with a 14-day quarantine for returning travellers who do not meet exemption criteria were introduced on 25^th^ March 2020 and these have remained in effect since.^4^ Localised lockdown policies have also been implemented, with variation in region-specific stay-at-home orders and, school closures.^5^

Previously, there was concern regarding the potential of increased healthcare burden from the dual impact of an ongoing COVID-19 pandemic in many countries coinciding with the seasonal influenza virus peak which causes significant annual morbidity and mortality.^6^ Understanding of the impact of COVID-19-related transmission mitigation measures on the transmission of other respiratory viruses is limited. Increasingly, it is being recognised that during the COVID-19 pandemic, the traditional respiratory virus season has been significantly altered with notable decreases in the incidence of other seasonal respiratory viral infections.^3,7,8^ The reasons for this observed significant decrease are not yet fully understood and are likely related, at least in part, to the aforementioned COVID-19 mitigation strategies.

The impact of the current COVID-19 pandemic on detection of influenza, respiratory syncytial virus and other seasonal respiratory viruses across all of Canada during the 2020-2021 influenza season has not previously been reported. The objective of this study is to characterise the epidemiology of influenza, respiratory syncytial virus, and other non-SARS-CoV-2 seasonal respiratory virus across Canada before and during the COVID-19 pandemic.

## Methods

### Design and setting

The study is a population-based observational study using Canada-wide laboratory surveillance data sources as detailed below.

### Data sources

Data on non-SARS-CoV-2 virus detection were obtained from the Canada Respiratory Virus Detection Surveillance System.^9^ This national surveillance is coordinated by the Public Health Agency of Canada (PHAC). Sentinel public health and hospital laboratories across Canada provide PHAC with weekly summaries of respiratory virus test results and test volumes. PHAC collates the data and delivers weekly publicly available updates. Reporting laboratories, include reference and public health laboratories for the Atlantic region, Province of Quebec, Province of Ontario, Prairies region, British Columbia and the Territories. For further details on laboratories included please refer to https://www.canada.ca/en/public-health/services/surveillance/respiratory-virus-detections-canada.html. Weekly Respiratory Virus Detections data are collected continuously, with reports available from 2006.

Data on SARS-CoV-2 cases in Canada were obtained from the Government of Canada Public Health Infobase.^10^ Public Health Infobase data is co-ordinated and managed by PHAC. The number of new daily cases of COVID-19 (confirmed and probable) are established from the net change between what provinces and territories report to PHAC for the current day and for the previous reported day.

Information on international and provincial travel restrictions and public health measures were obtained from the Government of Canada and Public Health Agency of Canada.^11^

All reported data in this study contains information licensed under the Open Government Licence – Canada. All data included in this analysis was obtained from publicly available de-identified datasets and therefore additional ethical approval was not required.

### Participants

The study population included all respiratory virus tests conducted at sentinel reporting laboratories in Canada during the study period of 2014 to 2021 inclusive. The reporting laboratories provide limited information on participant demographics and no clinical information on cases (either positive or negative) is available. Limited available demographic information was not analysed for this study.

### Measures/variables

In this study we analysed weekly data from the week ending 30^th^ August 2014 (epidemiological week 35) until the week ending the 13^th^ February 2021 (epidemiological week 6) inclusive. For the purpose of the study, we defined a case of non-SARS-CoV-2 respiratory virus as any laboratory-confirmed positive test for non-SARS-CoV-2 respiratory viruses reported to PHAC. Non-SARS-CoV-2 respiratory viruses included influenza, respiratory syncytial virus (RSV), parainfluenza viruses (PIV) types 1, 2, 3 and 4, adenovirus, human metapneumovirus (hMPV), enterovirus/rhinovirus, and seasonal coronaviruses. Seasonal coronavirus detection includes seasonal human coronaviruses HCoV-229E, HCoV-OC43, HCoV-NL63, HCoV-HKU1 and does not include human coronaviruses SARS-CoV, MERS-CoV, and SARS-CoV-2. Weekly percentage of tests positive for each virus was defined as the number of cases reported over the total number of tests reported for the epidemiologic week under surveillance, expressed as a percentage.

For SARS-CoV-2 cases we analysed daily data from 31^st^ January 2020 to 13^th^ February 2021 inclusive. For the purposes of this study, we defined weekly SARS-CoV-2 cases as the number of confirmed or probable cases of COVID-19 reported to PHAC for each week under study. For greater detail on case definition, fluctuation in daily case reporting across provinces and territories and a full list of web sources used in the epidemiological data please refer to health-infobase.canada.ca.^12^

### Statistical methods

The total study period included the 2014/2015 influenza season, defined as beginning at the week ending 30th August 2014 (epidemiological week 35) until the 2020/2021 influenza season, the week ending 13th February 2021 (epidemiological week 6). The baseline or control “pre-pandemic” period was defined as beginning from the week ending 30th August 2014 to the week ending 7th March 2020 inclusive. This period was selected to allow inclusion of five full seasons of baseline data until the global pandemic was called by the World Health Organization (WHO) on March 11, 2020.^13^ An inter-season period beginning at the onset of the COVID-19 pandemic was defined as the week beginning 14^th^ March 2020 (epidemiological week 11) until the week beginning 22^nd^ August 2020 (epidemiological week 11) inclusive and was excluded from the comparative analysis. The baseline period was compared to the current 2020/2021 influenza season to date, which we defined as the week ending 29^th^ August 2020 (epidemiological week 35) to the week ending the 13^th^ February 2021 (epidemiological week 6) inclusive.

Data analysis was performed using GraphPad Prism 9.0.2 (Graph-Pad Software, LLC) and Stata version 16.1. Interrupted time series analyses^14^ were performed to assess the difference in percent positivity for each virus of interest between the baseline pre-pandemic period and the current 2020/2021 influenza season period. Specifically, segmented negative binomial regression was used given the presence of over dispersion and each model included a) the number of positive tests; b) two pairs of Fourier sine-cosine terms; c) a nominal variable denoting the time period as either pre-pandemic, inter-season, and 2020/2021 influenza season; and d) an offset of the number of tests conducted. Rate ratios and 95% confidence intervals were generated between the pre-pandemic and 2020/2021 influenza season. A p-value of <0.05 was regarded as statistically significant.

The number of positive laboratory results was summarized by virus type using line graphs. Data on rhinovirus/enterovirus testing in the Province of Quebec, data on PIV, adenovirus, hMPV and rhinovirus testing in the Yukon territory and data on seasonal coronaviruses testing in Newfoundland were not available throughout the entire study period. One sentinel laboratory, St. Joseph’s Healthcare in Hamilton, Ontario, has been added to those providing surveillance data over the study period and began reporting data on the week ending 23^rd^ November 2019. Laboratories in the Territories region did not report data in the 2014/2015 season. One sentinel laboratory, Saskatoon laboratory, Saskatchewan, stopped reporting during the study period from the week ending 15^th^ February 2020. To identify the impact of these reporting differences, analysis was repeated excluding the data from St. Joseph’s Healthcare in Hamilton, the Territories and Saskatoon Laboratories.

This study is reported according to the STROBE guideline for observational studies.

## Results

A total of 6,722,859 tests for non-SARS-CoV-2 respiratory viruses (1,634,574 influenza; 541,699 enterovirus/rhinovirus; 1,468,261 RSV; 839,583 PIV; 607,797 seasonal coronaviruses; 793,990 hMPV, and 836,955 adenovirus) were recorded over the baseline pre-pandemic study period from sentinel laboratories across Canada. In the 2020/2021 season, a total of 1,101,553 tests for non-SARS-CoV-2 respiratory viruses were recorded from sentinel laboratories across Canada (292,003 influenza; 119,258 enterovirus/rhinovirus; 222,859 RSV; 122,381 PIV; 101,249 seasonal coronaviruses; 122,448 hMPV, and 121,355 adenovirus). The average weekly number of combined laboratory tests for all non-SARS-CoV-2 respiratory viruses performed was reported as 23,262 (range from 6,728 to 63,798) for the pre-pandemic period and 44,062 (range from 27,597 to 77,110) for the 2020/2021 season. Table 1 details average weekly testing for each virus of interest.

**Table 1.** Average weekly testing numbers and percentage positive tests for non-SARS-CoV-2 respiratory viruses at sentinel laboratories in Canada for the 2020/2021 influenza season and 2014-2019 pre-pandemic seasons. Baseline “Pre-pandemic” period begins the week ending 30^th^ August 2014 through to the week ending 7^th^ March 2020 inclusive. The 2020/2021 influenza season was defined as beginning the week ending 29^th^ August 2020 to week ending 13^th^ February 2021 inclusive. *Rate ratio and p-values calculated using interrupted time series to assess the difference in percent positivity for each virus of interest between the baseline pre-pandemic period and the current 2020/2021influenza season period. as described in methods. This table is based on weekly count data. Abbreviations: RSV = Respiratory syncytial virus, PIV = parainfluenza virus (includes combined type 1,2,3 and other types), hMPV = human metapneumovirus. ** Coronavirus excludes human coronaviruses SARS-CoV, MERS-CoV and SARS-CoV-2; Includes seasonal human coronaviruses HCoV-229E, HCoV-OC43, HCoV-NL63, HCoV-HKU1. Note data on rhinovirus/enterovirus testing in the Province of Quebec, data on PIV, adenovirus, hMPV and rhinovirus testing in the Yukon territory and data on seasonal coronaviruses testing in Newfoundland were not available for any week throughout the entire study period. One sentinel laboratory, St. Joseph’s Healthcare in Hamilton, Ontario, has been added to those providing surveillance data over the study period and began reporting data on the week ending 23^rd^ November 2019. Laboratories in the Prairies region did not report data in the 2014/2015 season. One sentinel laboratory, Saskatoon laboratory, Saskatchewan, stopped reporting during the study period from the week ending 15^th^ February 2020. Analysis excluding the data from St. Joseph’s Healthcare in Hamilton, the Territories and Saskatoon Laboratory did not appreciably change the results, with no change in p-values noted and minimal difference in magnitude any of the rate ratios (see table S1).

|  | **Pre-pandemic period**<br>(week ending 30th August 2014 - week ending 7th March 2020) |  | **2020/2021 influenza season**<br>(week ending 29th August 2020 - week ending the 13th February 2021) |  |  |  |
| --- | --- | --- | --- | --- | --- | --- |
| **Virus** | **Average weekly no. of laboratory tests (min-max)** | **Average weekly % positive tests (min-max)** | **Average weekly no. of laboratory tests (min-max)** | **Average weekly % positive tests (min-max)** | **Rate ratio of % positivity for 2020/2021 season versus pre-pandemic period (95% CI)\*** | **p-value\*** |
| Influenza A | 5529<br>(1251 - 17681) | 7.40<br>(0.11 - 33.97) | 11678<br>(4996 - 20715) | 0.013<br>(0 - 0.04) | 0.0017<br>(0.0011–0.0028) | <0.001 |
| Influenza B | 5529<br>(1251 - 17681) | 2.97<br>(0 - 17.02) | 11678 (4996 - 20715) | 0.007<br>(0 - 0.04) | 0.0061 (0.0030–0.0127) | <0.001 |
| RSV | 4973<br>(1204 - 16348) | 4.55<br>(0.11 - 17.80) | 8914 (4952 - 18413) | 0.051<br>(0.01 - 0.10) | 0.0145 (0.0105–0.0202) | <0.001 |
| PIV | 2856<br>(1027 - 7187) | 4.03<br>(1.15 - 9.74) | 4895 (3039 - 8486) | 0.059<br>(0 - 0.20) | 0.0172 (0.0127–0.0233) | <0.001 |
| Adenovirus | 2847<br>(1032 - 7207) | 2.04<br>(0.85 - 4.02) | 4854 (3039 – 7986) | 0.435<br>(0.19 - 0.82) | 0.2229 (0.1940–0.2562) | <0.001 |
| hMPV | 2698<br>(903 - 6890) | 2.51<br>(0 - 7.94) | 4891 (2917 - 8485) | 0.074<br>(0 - 0.19) | 0.0632 (0.0443–0.0902) | <0.001 |
| Enterovirus/<br>rhinovirus | 1838<br>(586 - 5980) | 17.73<br>(4.31 - 41.29) | 4770 (2914 - 8334) | 9.067<br>(3.56 - 24.12) | 0.5330 (0.4746–0.5977) | <0.001 |
| Coronaviruses** | 2066<br>(674 - 6413) | 2.72 (0 - 8.57) | 4050 (2265 - 6743) | 0.073<br>(0 - 0.26) | 0.0300 (0.0215–0.0418) | <0.001 |

By comparison, as of 13^th^ February 2021, 22,922,404 SARS-CoV-2 tests were reported to have been performed at reporting laboratories throughout Canada since the pandemic began.

We observed dramatically lower detection levels of all non-SARS-CoV-2 respiratory viruses during the 2020-2021 influenza season compared to baseline pre-pandemic levels (table 1, figure 1-3). This observed decrease did not seem to be related to a drop in overall numbers of tests for these viruses (supplemental figures S1 and S4). For influenza A and B, the percentage of tests with positive results during the 2020/2021 season decreased significantly, recorded at a rate of 0·0017 (95% CI: 0·001–0·003) and 0·0061 (95% CI: 0·003–0·013) times that of baseline, respectively. For RSV, the percentage of tests positive dropped to 0·0145 (95% CI: 0·011–0·020) times that of baseline during the 2020/2021season. For the remaining respiratory viruses studied, comparison of the percentage of tests positive for the 2020/2021 season with baseline also showed significant decreases. The percentage of tests with positive results for PIV decreased to 0·0017 (0·013–0·023) times that of baseline, for adenovirus this decreased to 0·223 (0·194–0·256) times that of baseline, for hMPV this decreased to 0·063 (0·044–0·090) times that of baseline, for enterovirus/rhinovirus this decreased to 0·533 (0·475–0.598) times that of baseline, and for seasonal coronaviruses this decreased to 0·030 (0·022–0·042) times that of baseline. Importantly, at the time of these observed decreases in percentage of positive tests for non-SARS-CoV-2 respiratory viruses, ongoing cases of SARS-CoV-2 were reported with an initial peak in cases in April 2020 followed by a larger second peak in January 2021 (Figure 1 b,c).

**Figure 1.**
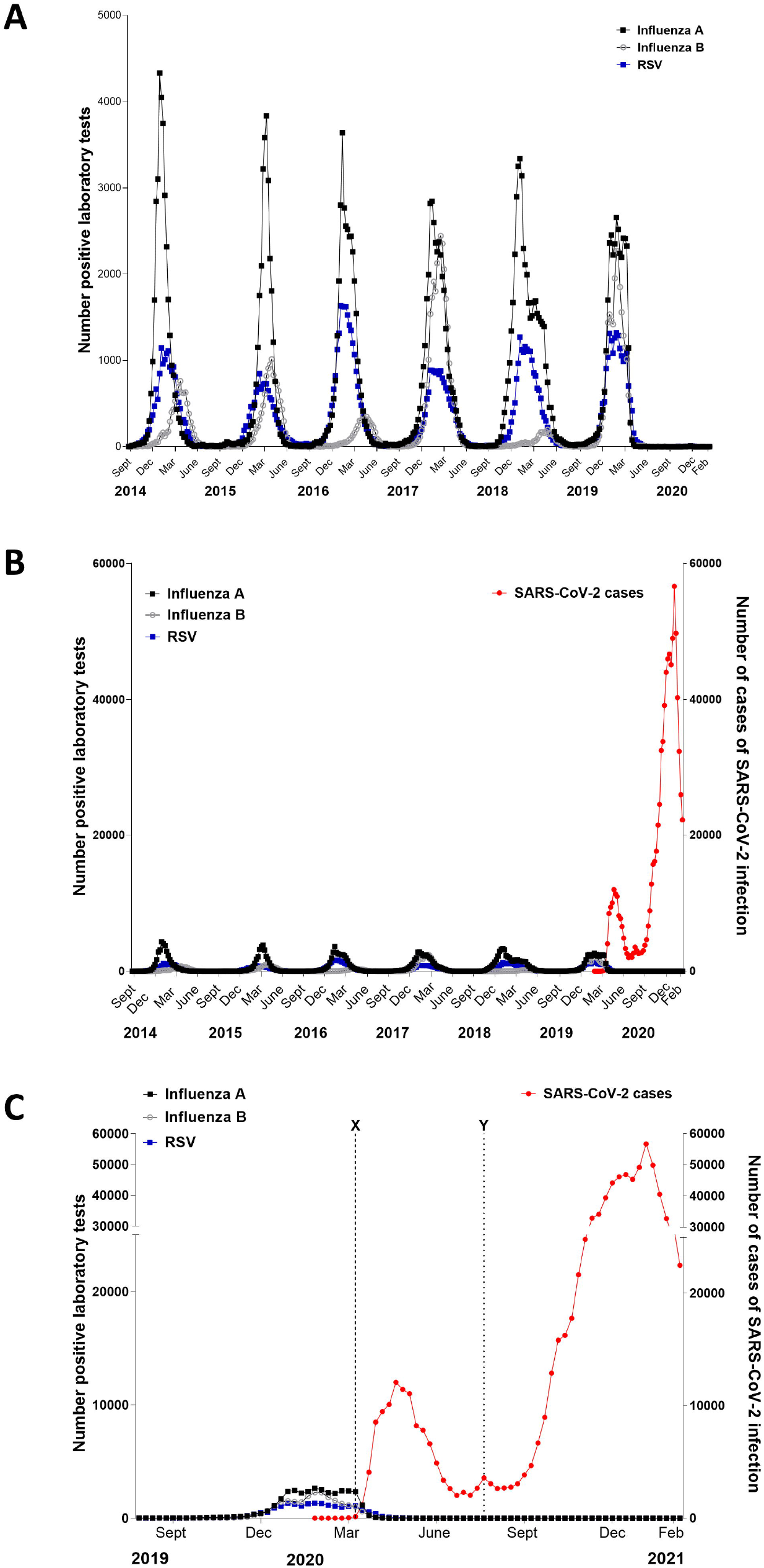
Seasonal variation in influenza and respiratory syncytial virus detection before and during the COVID-19 pandemic. For panels (A) and (B) dots represent individual weekly totals for all Canadian sites combined beginning the week ending 30th August 2014 to the week ending 13th February 2021 inclusive. Panel (A) shows the number of positive laboratory test results for influenza A, influenza B and RSV. Panel (B) shows the number of positive laboratory test results for influenza A, influenza B and RSV and the number of cases of COVID-19 (confirmed and probable). Panel (C) includes individual weekly totals for all Canadian sites combined for the week ending 3rd August 2019 until the week ending 13th February 2021. Canadian provincial and territorial governments declared emergencies from mid to late March 2020 and international travel restrictions were initiated 14th March 2020 (X). Canadian provincial regulations for face masks or face coverings in enclosed public spaces were introduced from July 7, 2020, beginning with Toronto and Ottawa (Y).^33^

**Figure 2.**
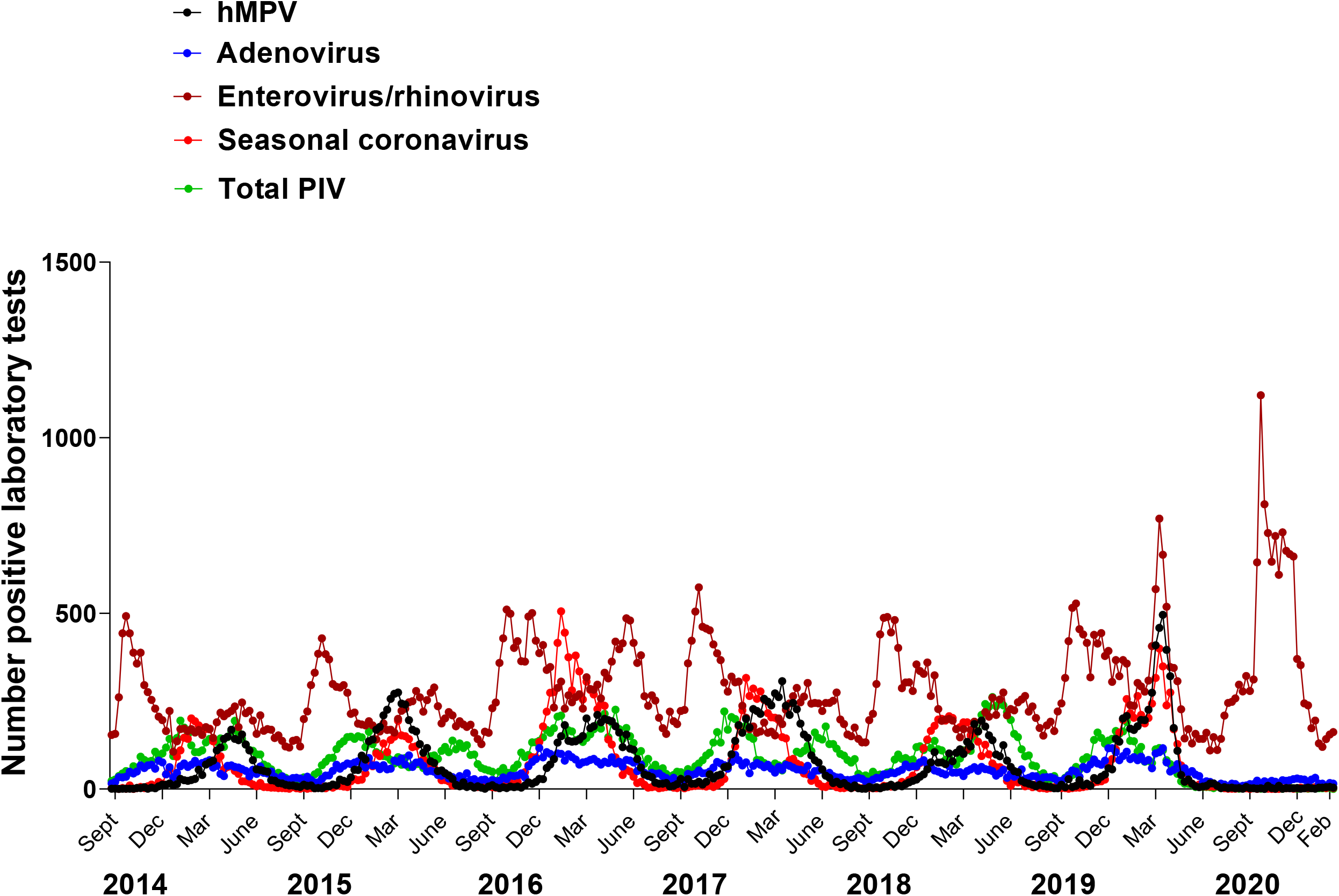
Seasonal variation in human metapneumonvirus (hMPV), adenovirus, enterovirus/rhinovirus, seasonal coronaviruses and parainfluenza viruses (PIV) before and during the COVID-19 pandemic. Graph shows the number of positive laboratory test results for hMPV, adenovirus, enterovirus/rhinovirus, seasonal coronaviruses and PIV. Dots represent individual weekly totals for the week ending 30th August 2014 to the week ending 13th February 2021 inclusive for all Canadian sites combined. PIV total includes numbers of positive tests for type 1,2,3 and 4. Coronavirus excludes human coronaviruses SARS-CoV, MERS-CoV and SARS-CoV-2; Includes seasonal human coronaviruses HCoV-229E, HCoV-OC43, HCoV-NL63, HCoV-HKU1.

**Figure 3.**
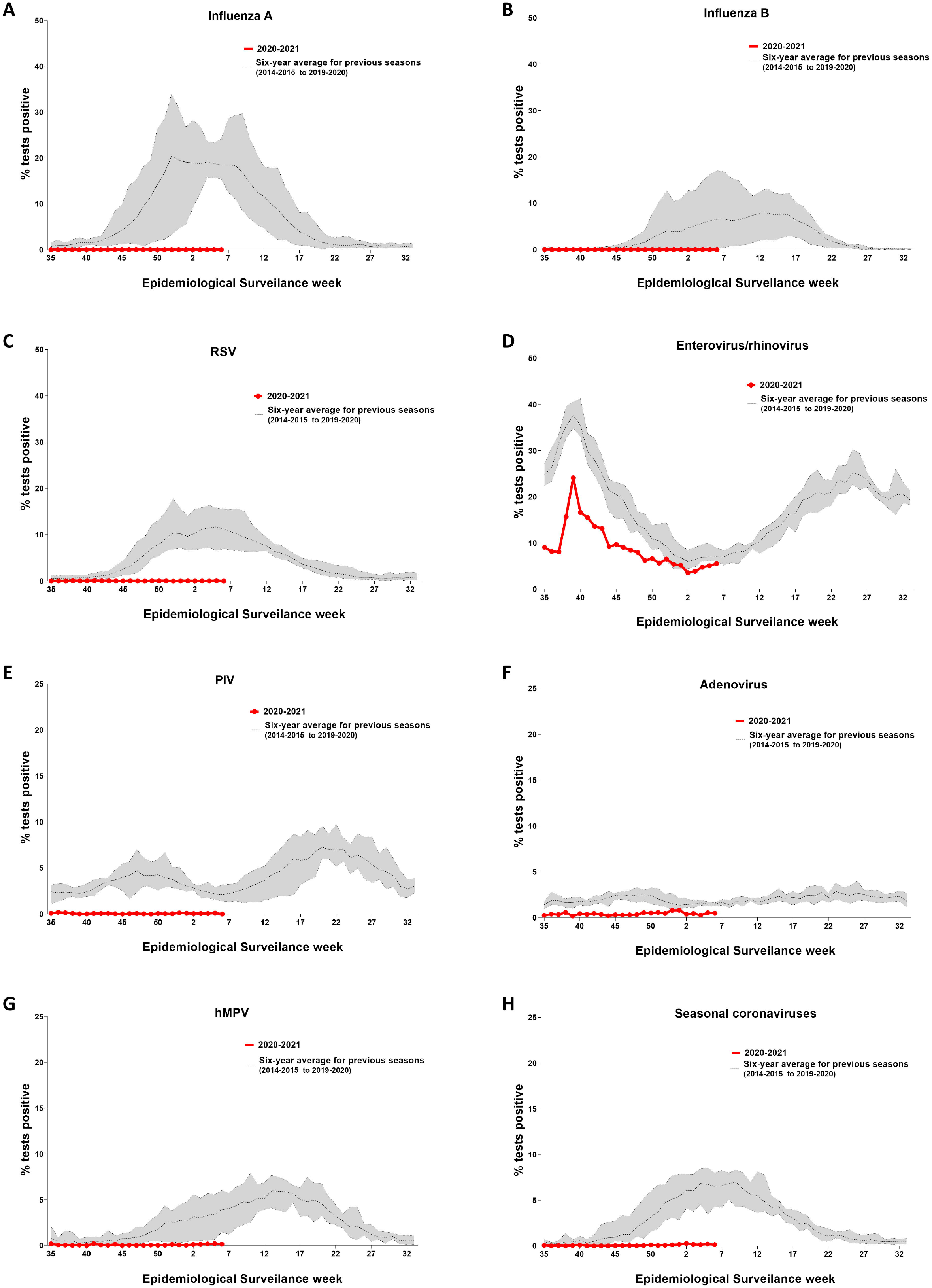
Temporal distribution of non-SARS-CoV-2 respiratory viruses by percentage test positivity in 2020/2021 season compared with previous years. Data plotted by epidemiological surveillance week. For 2020/2021 season data plotted from week 35 (week ending 29^th^ August 2020) to week 6 (week ending 13^th^ February 2021) 2020). The dotted line is the average percentage test positivity for the baseline seasons (from 2014-2015 to 2019-2020 influenza season). The shaded area represents the maximum and minimum percentage test positivity for the baseline seasons (from 2014-2015 to 2019-2020 influenza season). RSV respiratory syncytial virus, hMPV human metapneumovirus, PIV parainfluenza viruses (PIV includes numbers of positive tests for type 1,2,3 and 4). Coronavirus excludes human coronaviruses SARS-CoV, MERS-CoV and SARS-CoV-2; Includes seasonal human coronaviruses HCoV-229E, HCoV-OC43, HCoV-NL63, HCoV-HKU1.

Interestingly, higher (although still attenuated) ongoing detection of enterovirus/rhinovirus was reported during the 2020/2021 season versus other non-SARS-CoV-2 respiratory viruses, with average weekly percentage of tests positive of 9·1%, versus less than 1% respectively (table 1, figure 3a). On analysis by Canadian region, weekly percentage of tests positive for enterovirus/rhinovirus testing was relatively higher in the Territories (average 28·8, range: 11·75 - 47·5) and Atlantic region (average 19·0, range: 1·9 - 56·9), in comparison to the provinces of Ontario (average 7·7, range: 1·1 - 18·6), British Columbia (average 9·5, range: 2·0 - 23·7), and the region of the Prairies (average 7·5, range: 3·2 - 15·0) (see supplementary figure S5).

In a sensitivity analysis excluding the data from St. Joseph’s Healthcare in Hamilton, the Territories and Saskatoon Laboratory there was no appreciable change in the results, with no change in p-values noted and minimal difference in magnitude any of the rate ratios.

## Discussion

This study is the first Canada-wide study to examine the impact of the COVID-19 pandemic on the detection of both influenza and non-influenza seasonal respiratory viruses during the 2020-2021 influenza season. In this extensive study of public health and hospital microbiology laboratory data across Canada, we demonstrate a significant change in the seasonal variation of respiratory viral infections detected during the current COVID-19 pandemic. With the exception of ongoing enterovirus/rhinovirus detection, the usual seasonal peak in positive laboratory tests for common respiratory viruses, including non-SARS-CoV-2 coronaviruses was effectively absent during the 2020-2021 influenza season. This decrease in detection of non-SARS-CoV-2 respiratory viruses is consistent with studies from the United States and United Kingdom showing an early termination of the 2019/2020 influenza season and drop in detection of other viruses in the early phases of the pandemic.^3,15^ Similarly, a number of studies in Japan, Taiwan, Korea and Thailand demonstrated a marked decline in influenza virus detection in the early phases of the COVID-19 pandemic.^16–20^ As the pandemic has progressed and national lockdowns continued, studies of the respiratory virus season in the southern hemisphere from New Zealand and Australia demonstrated absence of the usual winter influenza virus seasonal peak and a marked reduction of other respiratory viruses.^21,22^

Typically in the Northern hemisphere, influenza, RSV and non-SARS-CoV-2 human coronaviruses are noted to peak during winter months, with adenovirus detection occurring throughout the year, peaks of parainfluenza viruses occurring in non-winter months and human metapneumoviruses and rhinoviruses being mainly detected during spring and fall seasons.^23^ The observed decrease in almost all seasonal respiratory viruses during the COVID-19 pandemic is likely related in large part to the implementation of multiple public health interventions, including restriction of international travel, hand-washing, wearing of face masks, school closures and stay-at-home orders. We note that at the same time of no detection of circulating seasonal coronaviruses, there were ongoing increases in cases of COVID-19 due to continuing SARS-CoV-2 transmission, which may reflect the difference in population susceptibility to the novel coronavirus. Indeed seasonal human coronavirus antibodies are not associated with protection against SARS-CoV-2 infection.^24^

We also reported continued detection of enterovirus/rhinoviruses during the 2020-2021 season. While this may suggest ongoing circulation of enterovirus/rhinoviruses despite public health measures to control COVID-19, variation in regional multi-layered public health measures may account for this continued circulation. Other studies have reported similar continued enterovirus/rhinovirus detections despite implementation of public health measures to reduce the spread of SARS-CoV-2. In a recent study of children in New South Wales, Australia, rhinovirus detections continued in the summer virus season of 2020 despite significant decreases in RSV detection over the same time period.^22^ Similarly, in a recent study in New Zealand, based on hospital-based severe acute respiratory surveillance data, between May and September 2020, marked reductions in rhinovirus levels were observed on the introduction of initial lockdown measures, however on easing restrictions a notable significant increase in rhinovirus-associated incidence rates was observed compared to other respiratory viruses.^21^ This potential for rhinovirus resurgence is important for the interpretation of syndromic surveillance models for COVID-19 as there is considerable overlap in clinical symptoms for even mild enterovirus/rhinovirus SARS-CoV-2.^25^

An apparent increase in rhinovirus detection on reduction of public isolation measures has also been reported in hospitalised adults in the UK following the re-opening of schools.^26^ Likewise, in Hong Kong, after the re-opening of schools and childcare centres, a large number of outbreaks of acute upper respiratory tract infections (URTIs), likely rhinovirus infections, were identified.^27^ The reasons for this reported resurgence of rhinovirus compared to other respiratory viruses are unclear. Rhinovirus is a non-enveloped virus that may be less susceptible to inactivation by handwashing.^22^ In addition, enteroviruses have the potential for fecal/oral transmission and it is possible that differences in the mechanism of enterovirus/rhinovirus transmission compared to other seasonal respiratory viruses mean more stringent public health measures are required to fully reduce transmission. On province-specific analysis of enterovirus/rhinovirus testing percentage positivity in Canada, regional variation during the 2020/2021 season was present. Higher percent positivity was seen in the Territories and Atlantic regions with reduced peaks compared to historical baseline observed for Ontario, British Columbia and The Prairies. This may reflect differences in regional implementation of public health measures, such as later introduction of compulsory facemask guidance, as well as differences in return to in-person classes for children and indoor gathering regulations.^28^ It is also possible that provincial/regional differences in SARS-CoV-2 activity may have played a role. In addition, rhinovirus infections are more likely to be symptomatic in children,^29^ who may be differentially impacted by regional variation in public health measures such as school closures and masking mandates. Due to the absence of age-specific data in this study we are unable to assess the role of circulating enterovirus/rhinovirus in the paediatric population specifically.

Overall, our results are consistent with a growing body of work indicating strict public health measures, such as regional lockdowns, border closures, handwashing and facemask wearing may be effective in significantly reducing the spread of epidemic respiratory viruses. However, what is notable in this study is the observed dramatic decrease in non-SARS-CoV-2 respiratory viruses including seasonal coronaviruses, despite ongoing detection of SARS-CoV-2 during the 2020/2021 winter season. This may reflect increased transmissibility of SARS-CoV-2 in comparison to other respiratory viruses, due to lack of preceding population immunity, such that it outcompetes seasonal respiratory viruses which are being more readily impacted by the measures implemented to mitigate SARS-CoV-2 transmission. It is difficult at this time to determine whether there is also potential impact of viral-viral interactions between SARS-CoV-2 and other respiratory viruses contributing to the observed decline in circulation. The potential for interaction between respiratory viruses has been speculated by previous modelling studies, such as work by Nickbakhsh *et al*. who found both positive and negative respiratory viruses interaction at population and individual host level and suggested that viral interference may account for reduced frequency of rhinovirus infections during influenza seasons.^30^ Nickbakhsh *et al*. further suggested such viral interference may potentially occur via interferon-mediated mechanisms. This concept is supported by recent *in vitro* work in differentiated airway epithelial cultures, demonstrating upregulation of interferon stimulated gene expression following rhinovirus infection, leading to significant inhibition of subsequent influenza A virus infection.^31^

Another potential explanation for the apparent dramatic decrease in seasonal respiratory viruses despite ongoing SARS-CoV-2 detection is a possible reduction in testing and/or laboratory reporting for non-SARS-CoV-2 respiratory viruses. We recognise one potential limitation of this study is that laboratories and hospitals may have become overburdened with implementing large scale testing for SARS-CoV-2 leading to changes in testing and reporting mechanisms for seasonal respiratory viruses compared to previous years. Although, importantly, we did not note a decrease in the total number of laboratory tests performed for each of the seasonal respiratory viruses studied during the pandemic compared to baseline. Indeed, early in the pandemic an apparent increase in the total number of tests performed for each virus of interest was observed (supplemental figure S4). However, it is likely that, compared to previous seasons, variation in testing practices by regional and local laboratories, such as increased periodic community testing, combined influenza/SARS-CoV-2 testing and increased testing for influenza and other seasonal respiratory viruses may have occurred during the 2020/2021 season. Moreover, it is worth noting that approaches for population testing of SARS-CoV-2 and non-SARS-CoV-2 viruses also differed, with testing of SARS-CoV-2 being much more community-based in comparison to other respiratory virus testing.

Finally, observed decreases in influenza virus detection may also be related, in part, to population changes in influenza vaccine uptake for the 2020/2021 season. On review of overall vaccine coverage data from the Canadian Seasonal Influenza Vaccination Coverage Survey,^32^ uptake by adults aged over 18 years in the 2020-2021 season was similar to the 2018-2019 and 2019-2020 seasons, but it is possible regional variation in uptake occurred.

## Limitations

There are a number of potential limitations of this study. Firstly, this is a retrospective observational study and it is difficult to identify whether the described decrease is as a direct result of multi-layered public health strategies to mitigate spread of SARS-CoV-2 or other unidentified factors. Strategies were added in close proximity and targeted specific populations, via for example school and workplace closures, at different times. These differing measures were implemented with substantial regional variation across Canadian provinces, such that identifying individual key factors contributing to the overall observed decrease in non-SARS-CoV-2 respiratory virus detection is not possible in this study.

This study used data from the Canadian Respiratory Virus Detections Surveillance System, which relies on reporting of results from individual laboratories. Changes in testing policies and eligibility at reporting laboratories may have led to differences in detection of non-SARS-CoV-2 respiratory viruses during the pandemic. Laboratory reporting remained consistent throughout the pandemic for most reporting laboratories and as detailed above. Analysis excluding data from sentinel laboratories with variation in reporting practice during the pandemic compared to baseline resulted in no significant change in the time-series or rate ratio results. Other changes in testing practices such as the introduction of COVID-19 assessment centres throughout Canada could have an impact on respiratory viruses detection during the pandemic. Furthermore, differences in health-seeking behaviour by individuals during the pandemic may also have altered the detection of non-SARS-CoV-2 respiratory viruses. For instance, in light of the concern for COVID-19, individuals with respiratory symptoms may have been more likely to attend for specific SARS-CoV-2 respiratory virus testing at designated testing centres rather than to receive testing for other respiratory viruses through usual mechanisms. Additionally, public health measures for the COVID-19 pandemic introduced asymptomatic screening and testing, as well as contact tracing and testing measures. This is likely to explain in part the higher numbers of SARS-CoV-2 tests and detections seen during the 2020-2021 respiratory virus season and may account for some of the differences observed in non-SARS-CoV-2 respiratory virus detection over the same period.

## Conclusion

In conclusion, in this study we report an effective absence of the annual seasonal epidemic for most non-SARS-CoV-2 respiratory viruses in Canada in 2020/2021 respiratory virus season. The reasons for this dramatic decrease are not yet clear and may reflect a number of different factors. It is likely the implementation of stringent measures to reduce the spread of SARS-CoV-2 also led to significant decreases in transmission of other respiratory viruses. However, the role of viral displacement and interference by SARS-CoV-2 is not yet known. In addition, the concept of a rebound in seasonal respiratory virus levels after relaxation of lockdown measures warrants further consideration. It is unclear whether the lack of exposure to non-SARS-CoV-2 respiratory viruses during the COVID-19 pandemic might result in larger outbreaks of other respiratory viral illnesses on easing of current public health measures in Canada. The absence of seasonal RSV and influenza epidemics also has potential implications for delivery of both palivizumab and influenza seasonal vaccination programs. Understanding the mechanisms behind the observed decreases in seasonal respiratory viruses is therefore of great importance and may benefit public health practice for mitigating seasonal respiratory virus epidemics and for informing responses to future respiratory virus pandemics.

## Data Availability

Anonymised raw data used to produce all the analyses, figures and tables for this paper are publically available for anyone who wishes to access the data. This data is available at the time of publication from the Government of Canada website via the links detailed in the methods section. Details of the conditions for use of this open dataset are available on the Government of Canada website.

## Data sharing statement

Anonymised raw data used to produce all the analyses, figures and tables for this paper are publically available for anyone who wishes to access the data. This data is available at the time of publication from the Government of Canada website via the links detailed in the methods section above. Details of the conditions for use of this open dataset are available on the Government of Canada website.

## Acknowledgements

The authors acknowledge the contribution of the provincial health and clinical laboratory partners that provide surveillance data to PHAC.

