## Supplemental figures and tables for "The impact of the COVID-19 pandemic on influenza, respiratory syncytial virus, and other seasonal respiratory virus circulation in Canada"

### Supplementary Figures and Tables

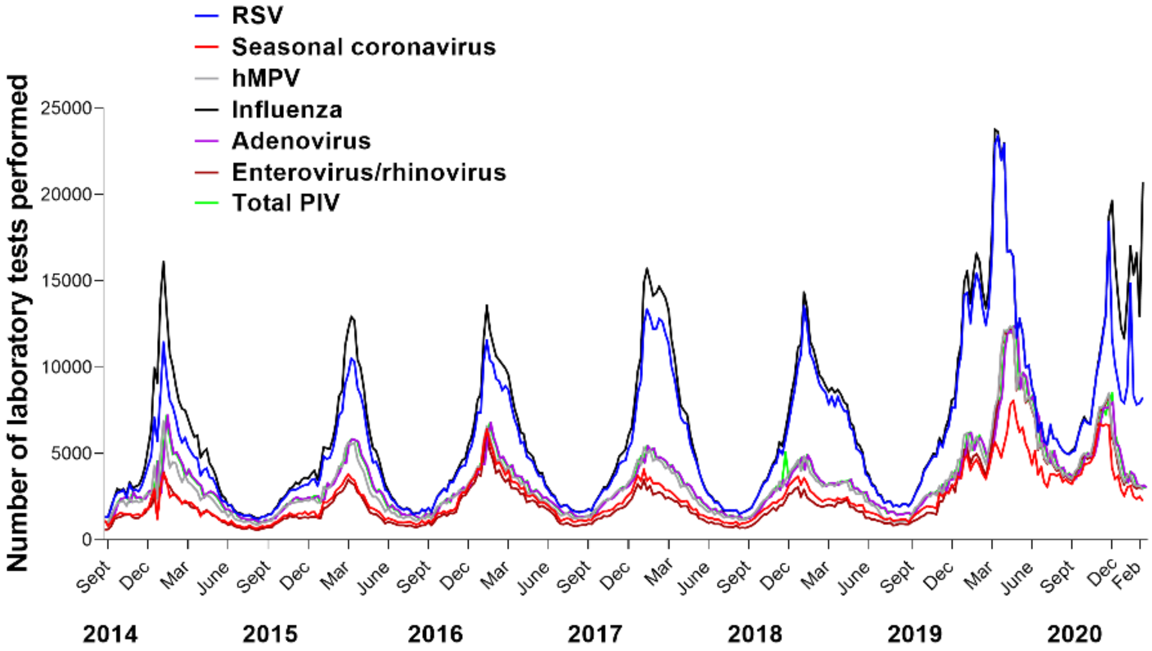


**Figure S1. The observed number of laboratory tests for combined Canada-wide data for all non-SARS-CoV-2 seasonal viruses reported.** Lines represent individual weekly totals for all Canadian sites combined for the week ending 30th August 2014 to the week ending 13th February 2021 inclusive. This includes; Influenza (all types combined), respiratory syncytial virus (RSV), human metapneumonvirus (hMPV), adenovirus, enterovirus/rhinovirus, seasonal coronaviruses and parainfluenza viruses (PIV). PIV total includes numbers of positive tests for type 1,2,3 and other types of PIV. Coronavirus excludes human coronaviruses SARS-CoV, MERS-CoV and SARS-CoV-2; Includes seasonal human coronaviruses HCoV-229E, HCoV-OC43, HCoV-NL63, HCoV-HKU1.

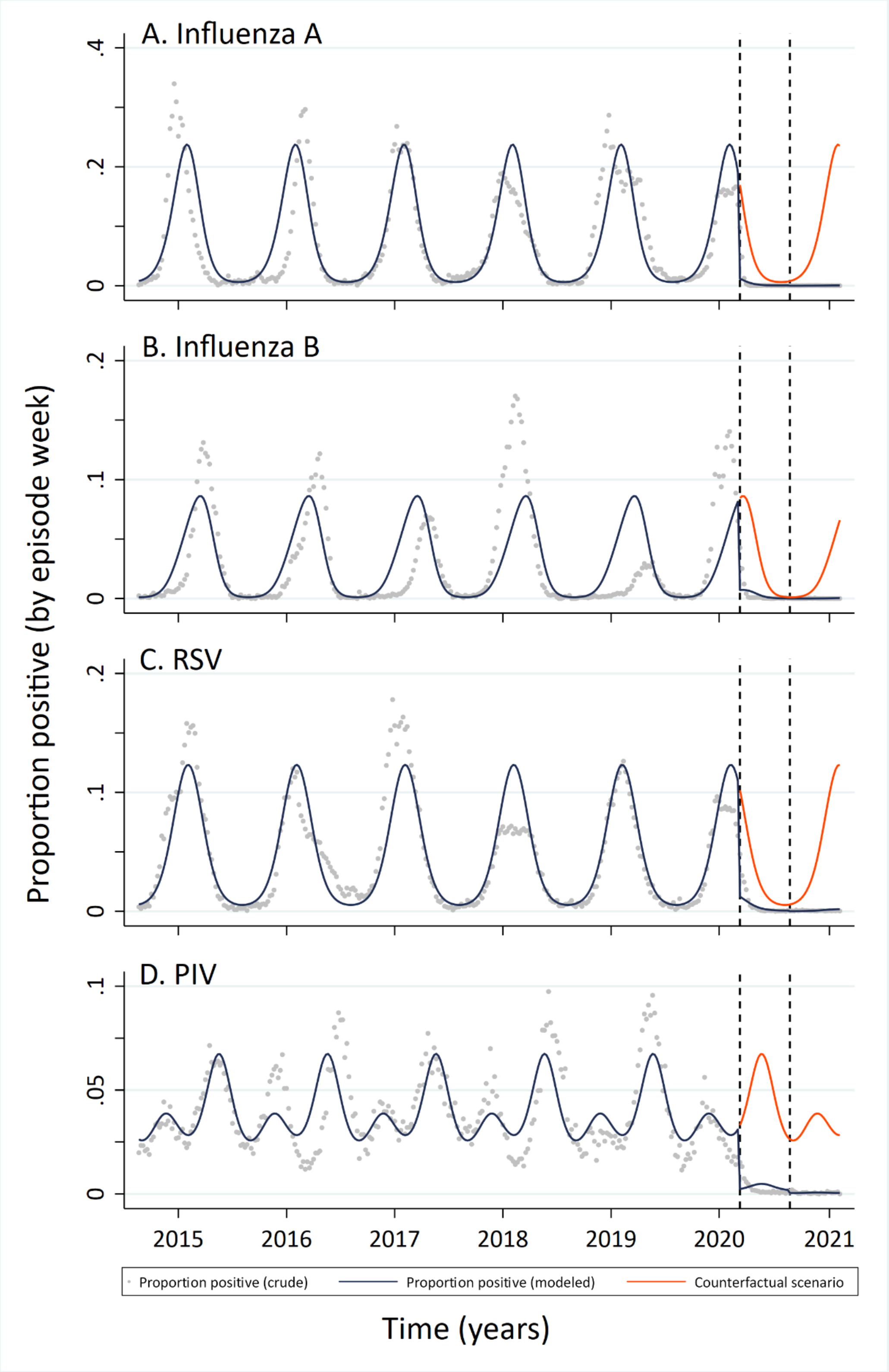


**Figure S2. Interrupted time series analyses of the proportion of positive laboratory tests for combined Canada-wide data for Influenza A, influenza B, Respiratory syncytial virus (RSV) and total parainfluenza viruses (PIV) for the week ending 30th August 2014 to the week ending 13th February 2021 inclusive.** Graphs demonstrate the actual (crude) proportion positive and modelled proportion positive for each season, including predicted admission for the 2020-2021 influenza season (counterfactual scenario).

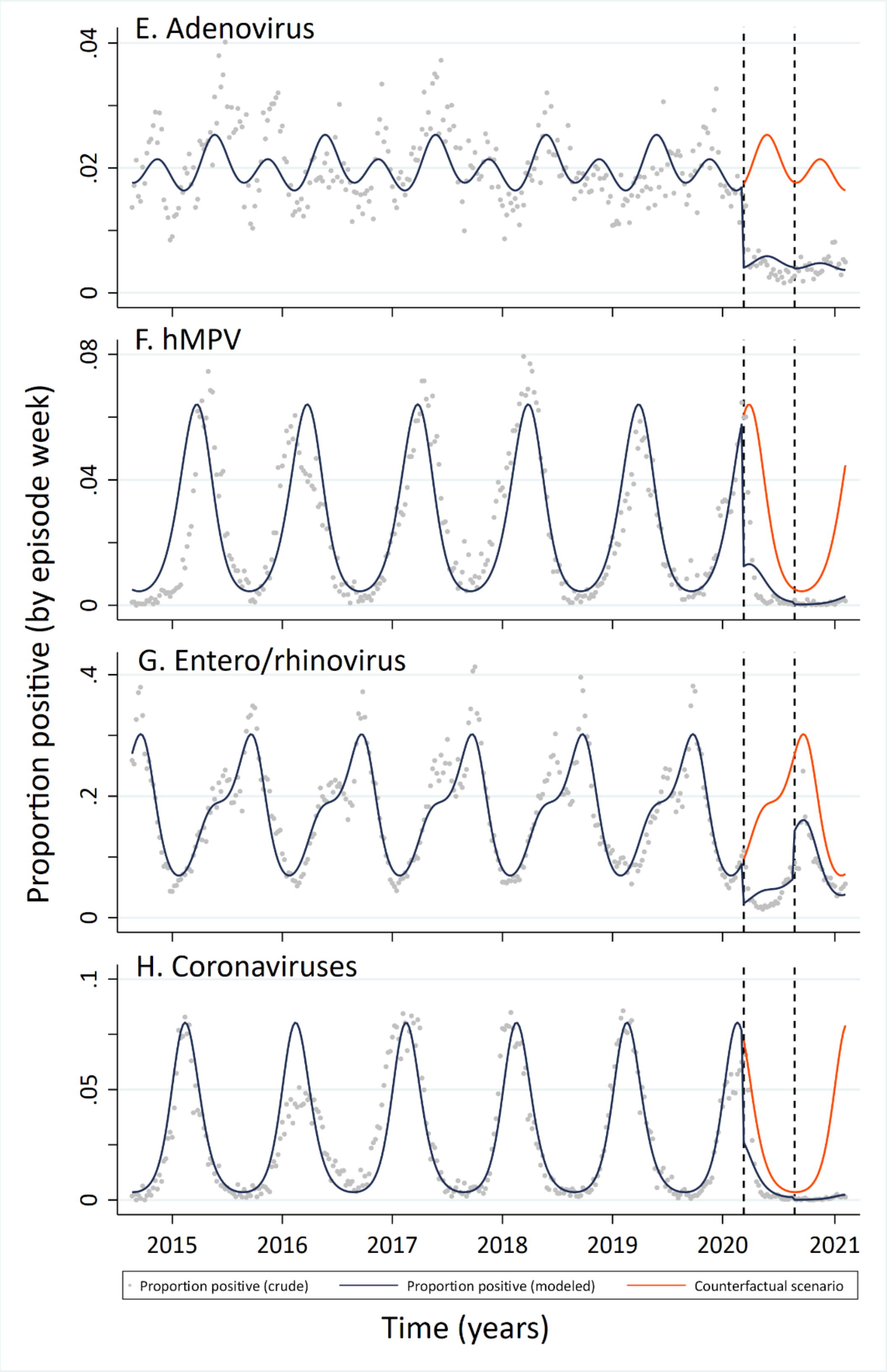


**Figure S3. Interrupted time series analyses of the proportion of positive laboratory tests for combined Canada-wide data fro adenovirus, human metapneumonvirus (hMPV), enterovirus/rhinovirus and seasonal coronaviruses (excludes SARS-CoV-2) for the week ending 30th August 2014 to the week ending 13th February 2021 inclusive.** Graphs demonstrate the actual (crude) proportion positive and modelled proportion positive for each season, including predicted admission for the 2020-2021 influenza season (counterfactual scenario).

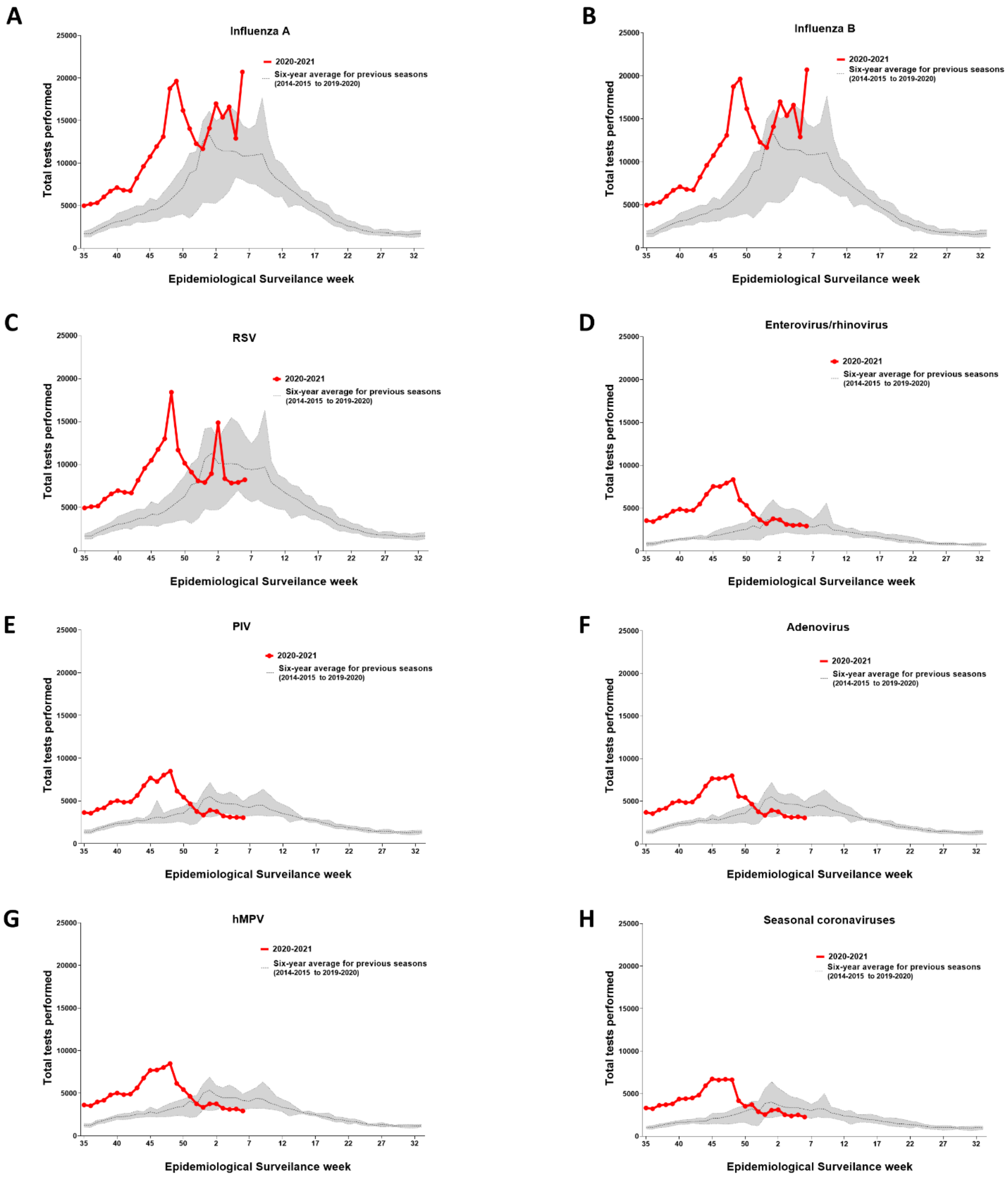


**Figure S4. Temporal distribution of testing for non-SARS-CoV-2 respiratory viruses in 2020/2021 season compared with previous years.** Data plotted by epidemiological surveillance week. For 2020/2021 season data plotted from week 35 (week ending 29th August 2020) to week 6 (week ending 13th February 2021) 2020). The dotted line is the average number of tests reported for the baseline seasons (from 2014-2015 to 2019-2020 influenza season). The shaded area represents the maximum and minimum number of respiratory virus tests reported by week during the 2014-2015 to 2019-2020 influenza seasons. RSV respiratory syncytial virus, hMPV human metapneumovirus, PIV parainfluenza viruses (PIV includes numbers of positive tests for type 1,2,3 and 4). Coronavirus excludes human coronaviruses SARS-CoV, MERS-CoV and SARS-CoV-2; Includes seasonal human coronaviruses HCoV-229E, HCoV-OC43, HCoV-NL63, HCoV-HKU1.

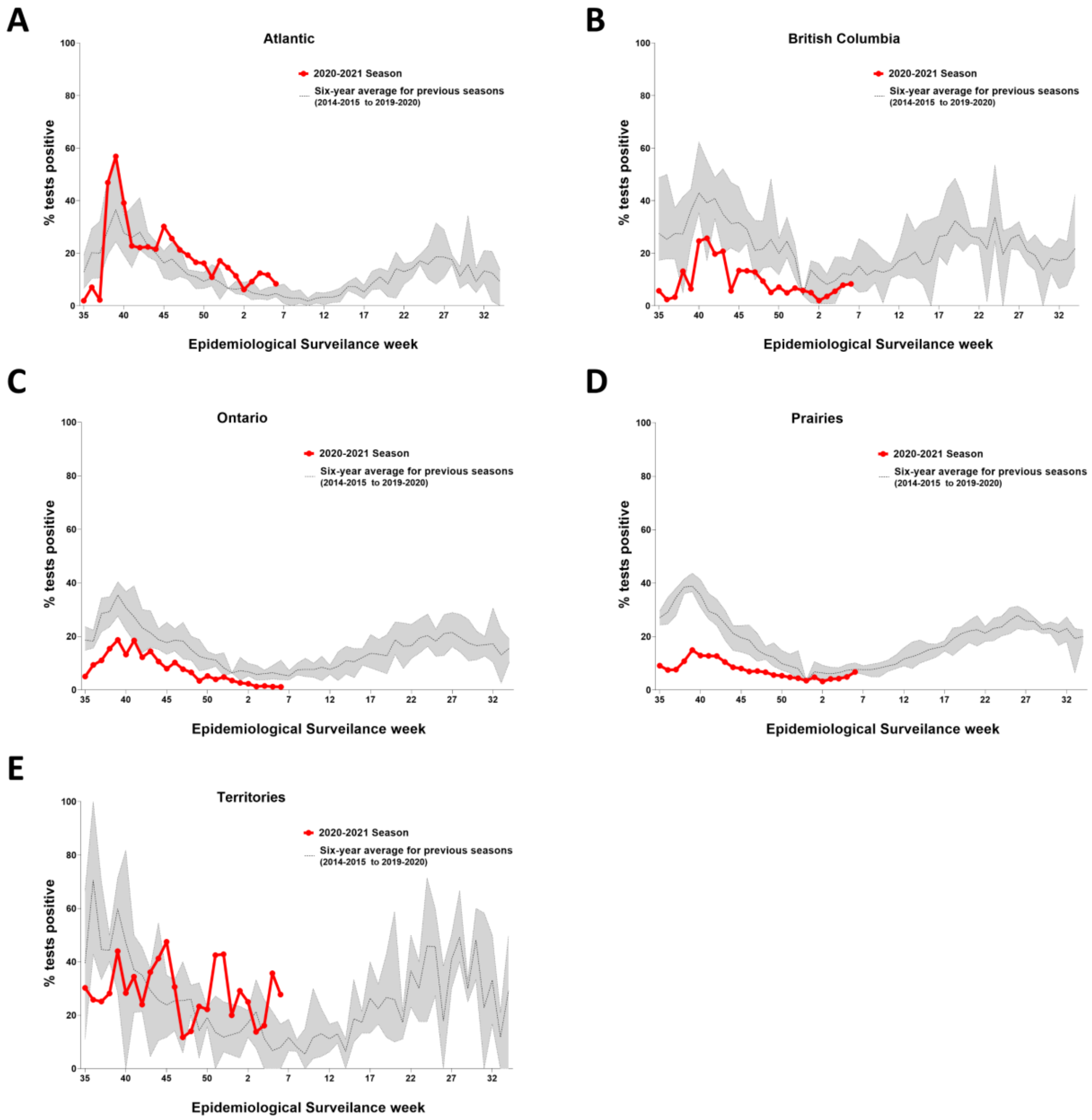


**Figure S5. Temporal distribution of percentage positive tests for enterovirus/rhinovirus by Canadian province for the 2020/2021 season compared with baseline.** Data plotted by epidemiological surveillance week. For 2020/2021 season data is plotted from week 35 (week ending 29th August 2020) to week 6 (week ending 13th February 2021) inclusive. The dotted line is the average percent positive test results at baseline (2014-2015 to 2019-2020 influenza seasons). The shaded area represents the maximum and minimum positive test results at baseline (2014-2015 to 2019-2020 influenza seasons). Data for the Province of Québec was not available.

**Table S1.** **Rate ratios for difference in percent positivity for non-SARS-Co-V 2 respiratory viruses in Canada between baseline pre-pandemic and 2020/2021 influenza seasons showing secondary analysis to exclude laboratories with missing data.**

| Virus | Primary Analysis | | Sensitivity Analysis | |
|---|---|---|---|---|
| | Rate ratio vs. pre-pandemic period (95% CI) | p-value | Rate ratio vs. pre-pandemic period (95% CI) | p-value |
| Influenza A | 0.0017 (0.0011–0.0028) | <0.001 | 0.0013 (0.0008–0.0022) | <0.001 |
| Influenza B | 0.0061 (0.0030–0.0127) | <0.001 | 0.0029 (0.0013–0.0066) | <0.001 |
| RSV | 0.0145 (0.0105–0.0202) | <0.001 | 0.0139 (0.0101–0.0191) | <0.001 |
| PIV | 0.0172 (0.0127–0.0233) | <0.001 | 0.0169 (0.0124–0.0232) | <0.001 |
| Adenovirus | 0.2229 (0.1940–0.2562) | <0.001 | 0.2218 (0.1920–0.2564) | <0.001 |
| hMPV | 0.0632 (0.0443–0.0902) | <0.001 | 0.0612 (0.0420–0.0892) | <0.001 |
| Entero/rhinovirus | 0.5330 (0.4746–0.5977) | <0.001 | 0.5177 (0.4605–0.5820) | <0.001 |
| Coronaviruses | 0.0300 (0.0215–0.0418) | <0.001 | 0.0298 (0.0213–0.0418) | <0.001 |

Rate ratio and p-values calculated using interrupted time series to assess the difference in percent positivity for each virus of interest between the baseline pre-pandemic period and the current 2020/2021influenza season period as described. Primary analysis includes all sentinel laboratory data. Secondary (sensitivity) analysis was repeated excluding the data from St. Joseph's Healthcare in Hamilton, the Territories and Saskatoon Laboratory. Abbreviations: RSV = Respiratory syncytial virus, PIV = parainfluenza virus (includes combined type 1,2,3 and other types), hMPV = human metapneumovirus. Coronavirus excludes human coronaviruses SARS-CoV, MERS-CoV and SARS-CoV-2; Includes seasonal human coronaviruses HCoV-229E, HCoV-OC43, HCoV-NL63, HCoV-HKU1.
